# Endoscopic score predicting recurrence after foam sclerotherapy in grade I internal hemorrhoids: Development and external validation

**DOI:** 10.1101/2025.10.28.25338987

**Authors:** Fei-Yu Zhang, Lei-Ming Xu, Feng-Yu Gao, Wen Wang, Wu-Lian Lin, Hao Zhang, Ke-Zhou Wang, Qing-Jing Wang, Ao-Jia Jin, Ru-Ping Yang, Chun-Ying Qu, Yi Zhang, Zheng-Hong Li, Dou Wang, Cui-Cui Shi, Tian-Ben Shen, Fei Shen, Ye Hu, Feng Shen

## Abstract

**BACKGROUND:** Foam sclerotherapy is an effective minimally invasive treatment for grade I internal hemorrhoids, but recurrence limits long-term benefit and no validated endoscopic tool is available for individualized risk prediction.

**AIM:** To develop and validate an endoscopy-based score for predicting recurrence after foam sclerotherapy in patients with grade I internal hemorrhoids.

**METHODS:** In this prospective, multicenter observational cohort, consecutive adults with grade I internal hemorrhoids treated with 1% polidocanol foam were enrolled. A Cox model was developed from clinical and standardized endoscopic predictors. Discrimination, calibration, bootstrap internal validation, and 36-month decision curve analysis were assessed. The unchanged model was evaluated in an independent external cohort, and a separate cohort provided exploratory endoscopic and histopathological biological verification.

**RESULTS:** A total of 483 patients were included in model development; 115 developed recurrences during follow-up, with cumulative recurrence rates of 10.1% at 24 months and 20.0% at 36 months. The weighted continuous Endoscopic Hemorrhoid Recurrence Score was: Endo-HRS = 1 × male sex + 4 × number of hemorrhoids + 11 × maximum hemorrhoid diameter (cm) + 2 × red color sign grade (0–3). A nomogram estimated individual 36-month recurrence probability. Discrimination was good in development (C-index 0.82; continuous-model 36-month area under the curve 0.88) and external validation (*n* = 279; bootstrap-corrected C-index 0.90, 95%CI: 0.862–0.933). Exploratory endoscopic and histopathological findings were directionally consistent with higher-risk groups.

**CONCLUSION:** Endo-HRS is a weighted endoscopy-based recurrence score with encouraging internal and external performance. It may support risk stratification and inform future evaluation of risk-adapted follow-up, but its clinical impact requires prospective assessment.

Core Tip: In this prospective multicenter study, we developed and externally evaluated Endo-HRS, a weighted continuous score based on sex, number of hemorrhoids, maximum hemorrhoid diameter, and red color sign grade. The model showed good discrimination in the development and external cohorts, and a nomogram estimates individual 36-month recurrence probability. Exploratory endoscopic and histopathological findings supported biological plausibility. Endo-HRS may aid risk stratification, although prospective impact studies are required before it is used to direct surveillance or retreatment.

**Graphical Abstract:** Patients with Goligher grade I internal hemorrhoids receiving foam sclerotherapy were evaluated using standardized clinical and endoscopic features. Endo-HRS was developed and externally evaluated, with exploratory endoscopic and histopathological biological verification. The score provides recurrence-risk stratification and may inform the design of future risk-adapted follow-up studies. By Figdraw. (ID: ORWRTaae33)

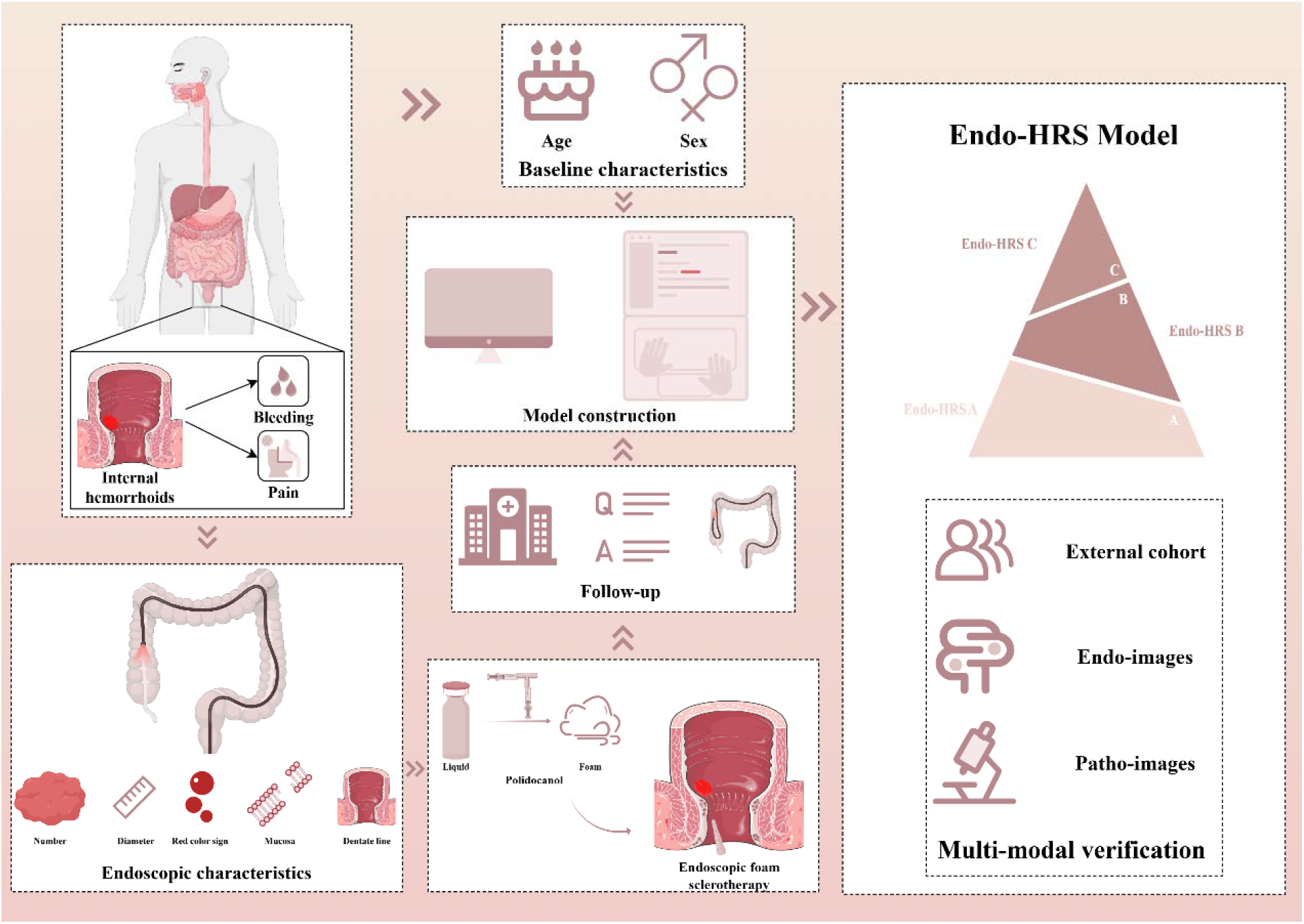

## INTRODUCTION

Internal hemorrhoids, characterized by symptomatic enlargement or distal displacement of the anal vascular cushions, are among the most common anorectal disorders. The main clinical manifestations are bleeding, prolapse, and discomfort[1, 2]. Available treatments range from conservative measures to definitive surgical intervention[3, 4]. Among minimally invasive options, endoscopic foam sclerotherapy with polidocanol has attracted growing interest because of its procedural simplicity, rapid recovery, and favorable safety profile[5–7]. However, recurrence remains a major limitation[8, 9], and currently available clinical grading systems do not provide a robust framework for individualized recurrence prediction.

Endoscopic polidocanol-based therapies have recently emerged as effective minimally invasive approaches for internal hemorrhoids[10, 11]. A multicenter randomized study demonstrated favorable efficacy and safety, with rapid recovery and low complication rates[12–14]. Nevertheless, recurrence after treatment remains an unresolved problem, and reliable tools for individualized risk stratification are lacking.

Conventional grading does not fully characterize the endoscopic phenotype of hemorrhoidal disease[15, 16]. The Goligher classification is driven primarily by prolapse and has recognized limitations for clinical research, whereas pathophysiological and symptom-oriented frameworks remain mainly descriptive[17–21]. Standardized endoscopic assessment of hemorrhoid number, size, surface vascular changes, mucosal erosion, and dentate-line injury may therefore provide more objective information for recurrence modeling[22, 23].

Therefore, we conducted a prospective multicenter observational study to develop and externally validate a novel endoscopic model for predicting recurrence after foam sclerotherapy in patients with grade I internal hemorrhoids.

## MATERIALS AND METHODS

### Study design and population

A prospective, multicenter, observational cohort study was conducted to develop and externally validate an endoscopic prediction model for recurrence after foam sclerotherapy for grade I internal hemorrhoids, in line with the STROBE reporting framework[24]. Consecutive patients were enrolled between January 2018 and December 2022 at four tertiary gastroenterology and endoscopy centers in China: Xinhua Hospital, Shandong Provincial Maternal and Child Health Care Hospital, the 900th Hospital of the PLA Joint Service Support Force, and Baoshan People’s Hospital. The target sample size was determined according to the expected number of recurrence events required for multivariable time-to-event modeling. Assuming a 3-year recurrence rate of approximately 20%-30%, a target enrollment exceeding 400 patients was prespecified.

Eligible participants were adults aged 18–70 years with endoscopically confirmed Goligher grade I internal hemorrhoids who underwent 1% (20 mg/2 mL) polidocanol (Aethoxysklerol; Hameln Pharmaceuticals, Germany) foam sclerotherapy. Key exclusion criteria included grade II–IV or external hemorrhoids, concomitant anorectal or perianal disease, contraindications to sclerotherapy, and other protocol-defined ineligibility criteria. Detailed inclusion and exclusion criteria are provided in Supplementary Tables S1 and S2. Chronic constipation or straining, prolonged sitting or standing, alcohol and spicy-food exposure, and pregnancy or childbirth history were not prospectively collected using standardized definitions and therefore were not entered into the primary model.

All participants provided written informed consent in accordance with the Declaration of Helsinki[25]. The study was prospectively registered at ClinicalTrials.gov (NCT04398823).

### Endoscopic therapy and data collection

Patients followed a low-fiber diet on the day before colonoscopy and underwent split-dose bowel preparation with polyethylene glycol solution (Hengkangzhengqing; Hygecon, Shangrao, Jiangxi, China) according to ESGE recommendations[26]. Foam sclerotherapy was performed using a standardized technique[12]. Specifically, 1% polidocanol was prepared using Tessari’s method[27] and injected through a 25-gauge InterjectTM needle (Boston Scientific, Marlborough, MA, United States) under endoscopic visualization (H290 series, Olympus, Tokyo, Japan).

Two experienced endoscopists completed standardized training and independently recorded the prespecified baseline endoscopic variables before consensus resolution. These variables included number of hemorrhoids, maximum hemorrhoid diameter, red color sign grade, mucosal erosion, and dentate-line injury. The evaluators were blinded to the treating operators. Discrepancies were resolved by consensus, and the final consensus values were used for analysis. Although endoscopic findings were independently assessed and disagreements were resolved by consensus, the individual pre-consensus ratings were not retained in the final study database. Consequently, formal interobserver agreement statistics could not be reconstructed. Number of hemorrhoids was confirmed using standardized antegrade and retroflexed views[22, 23]. Maximum hemorrhoid diameter was measured with an endoscopic scale marker. The red color sign was graded from 0 to 3 according to vascular congestion: 0, absent; 1, scattered red dots on the hemorrhoid surface; 2, linear red streaks; and 3, a large confluent area of congestion (Supplementary Figure S1). Mucosal erosion was recorded as present or absent. Dentate-line injury was classified as 0 (none), 1 (< 0.5 cm), or 2 (≥ 0.5 cm).

### Follow-up and outcomes

All patients underwent a minimum follow-up of 6 months, with scheduled clinic visits at 3-month intervals. Each visit included a clinical interview, digital rectal examination, and standardized questionnaire assessment (Supplementary Table S3). Protocolized surveillance colonoscopy was performed every 6 months irrespective of symptoms. Endoscopic confirmation was mandatory for suspected recurrence, and all re-examination records were collected prospectively.

Time to recurrence was recorded in months. Thirty-six months was the prespecified primary prediction horizon rather than the maximum observation period. Participants were followed until confirmed recurrence, last documented contact, or administrative censoring on December 31, 2025; non-recurrent participants could be observed beyond 36 months. The maximum development-cohort follow-up was 87 months. The primary endpoint was endoscopically confirmed reappearance of internal hemorrhoids associated with bleeding or definite hemorrhoidal mucosal vascular abnormality during scheduled surveillance or symptom-prompted examination. Outcome adjudicators were blinded to baseline Endo-HRS predictors and risk groups, and disagreements were resolved by independent review. Symptom-only episodes without objective recurrent hemorrhoids were recorded separately and were not counted as primary events because bleeding and anorectal discomfort may arise from other causes. To evaluate the potential conceptual overlap between baseline red color sign grade and recurrence assessment, a prespecified sensitivity analysis was performed by refitting the multivariable Cox model after excluding baseline red color sign grade from the candidate predictors.

### Model development

Candidate predictors were prespecified according to clinical and endoscopic relevance and then summarized using univariable Cox proportional hazards models. Variables with P < 0.10 and prespecified clinically relevant variables were considered in multivariable Cox modeling. Backward selection was used for the final model, and its potential for model instability and optimism is acknowledged. The proportional hazards assumption was assessed using Schoenfeld residuals. The functional form of maximum hemorrhoid diameter was assessed using restricted cubic splines, with nonlinearity-test result 0.638. Full regression coefficients, standard errors, hazard ratios, 95%CIs, *P* values, and baseline survival estimates are reported in Supplementary Table S4.

The Endoscopic Hemorrhoid Recurrence Score (Endo-HRS) was constructed as a weighted continuous risk score derived from the rescaled Cox linear predictor; decimal values are possible because maximum hemorrhoid diameter is entered in centimeters. Model discrimination was assessed using Harrell’s concordance index and time-dependent receiver operating characteristic curves. Calibration was assessed using plots, calibration slope, integrated Brier score (IBS), and the 36-month Brier score. Internal validation used 1000 bootstrap resamples to estimate optimism and optimism-corrected performance. Clinical utility at 36 months was evaluated by decision curve analysis against treat-all and treat-none strategies[28, 29]. Apparent, internal, and external performance metrics are summarized in Supplementary Table S5.

Thresholds were derived from the 36-month time-dependent receiver operating characteristic analysis. The lower rule-out threshold and upper rule-in threshold defined the three-tier classification: group A, < 18.2; group B, 18.2 to < 27.4; and group C, ≥ 27.4. The Youden-optimized threshold of 22.8 is reported as a separate binary reference and was not the sole basis for the three-tier grouping. Recurrence-free interval among groups was compared using Kaplan-Meier analysis.

### Web-based Endo-HRS calculator

A web-based calculator was implemented to generate the weighted Endo-HRS and classify patients into groups A, B, and C. The calculator is available at https://feiyuzhang.shinyapps.io/EndoHRS-Calculator/. The website is an optional implementation rather than the sole method of calculation; the complete formula and a printable worksheet are provided in Supplementary Table S6. The corresponding authors plan to maintain the web application for every six months and will provide source code or an offline calculator upon reasonable request if the website becomes unavailable.

### External cohort validation of the Endo-HRS model

The external cohort included 291 patients screened between January and December 2022 at five contributing external centers: The Ninth People’s Hospital of Shanghai Jiao Tong University School of Medicine, Shanghai Tongren Hospital, Shanghai Construction Engineering Hospital, The Fifth People’s Hospital of Ganzhou City, Jiangxi Province, and Shigatse People’s Hospital. This constituted geographic validation. The same predictor and outcome definitions were used, and no external-cohort data were used for model development, coefficient estimation, or refitting. The external cohort included 60 recurrence events, with median potential follow-up of 33.2 months (95% CI 32.4-34.1), administrative censoring on December 31, 2025, and maximum follow-up of 48 months. Baseline characteristics are presented in Supplementary Table S7.

The original Cox model was applied to the external cohort without refitting. Individual linear predictors and absolute risks were calculated using the development-cohort coefficients and baseline survival. Discrimination was evaluated using the C-index with 95%CI from 1000 bootstrap resamples and time-dependent receiver operating characteristic analysis at 24, 36, and 48 months. Calibration at 36 months was assessed using grouped estimates, logit-LOESS smoothing, calibration intercept, calibration slope, and Brier score. Decision curve analysis was also evaluated at the 36-month horizon.

### Exploratory multimodal biological verification

A separate exploratory biological-verification cohort comprised 40 participants who were not included in the development or external validation cohorts. All 40 contributed standardized antegrade and retroflexed endoscopic images. Two blinded endoscopists independently reviewed the images, with disagreements resolved by a third reviewer. Representative hematoxylin-eosin and Masson-stained sections were examined; quantitative collagen-proportionate-area analysis was restricted to 15 adequate specimens, with 5 specimens in each Endo-HRS group. This component was designed to explore biological plausibility rather than to validate predictive performance or establish causality.

Hemorrhoid tissue biopsies were obtained using standard forceps and processed for hematoxylin-eosin and Masson’s trichrome staining to characterize venous congestion and collagen deposition. Microvascular features and collagen-proportionate area were quantified using ImageJ software[30].

### Statistical analysis

Continuous variables were summarized as mean ± SD or median (interquartile range), as specified, and categorical variables as frequencies (%). Group comparisons used the χ2 test, Wilcoxon rank-sum test, or other appropriate methods. Age was reported consistently as median (interquartile range).

All baseline predictor variables were completely recorded, and therefore no imputation was performed. The nine development-cohort and seven external-cohort participants lost before completion of the minimum follow-up were excluded from the primary time-to-event analyses and are shown in Figure 1. Complete-case analyses were used as sensitivity analyses. Median potential follow-up was estimated using the reverse Kaplan-Meier method, and censoring distributions were examined. Sensitivity analyses assessed the stricter bleeding-confirmed recurrence endpoint, exclusion of sex from the model, and complete-case versus imputed estimates. All analyses were performed in R (version 4.5.1; R Foundation for Statistical Computing, Vienna, Austria) and SPSS (version 27.0; IBM Corp., Armonk, NY, United States). Relevant R packages are listed in Supplementary Table S8. Custom scripts are available from the corresponding authors upon reasonable request.

**Figure 1.**
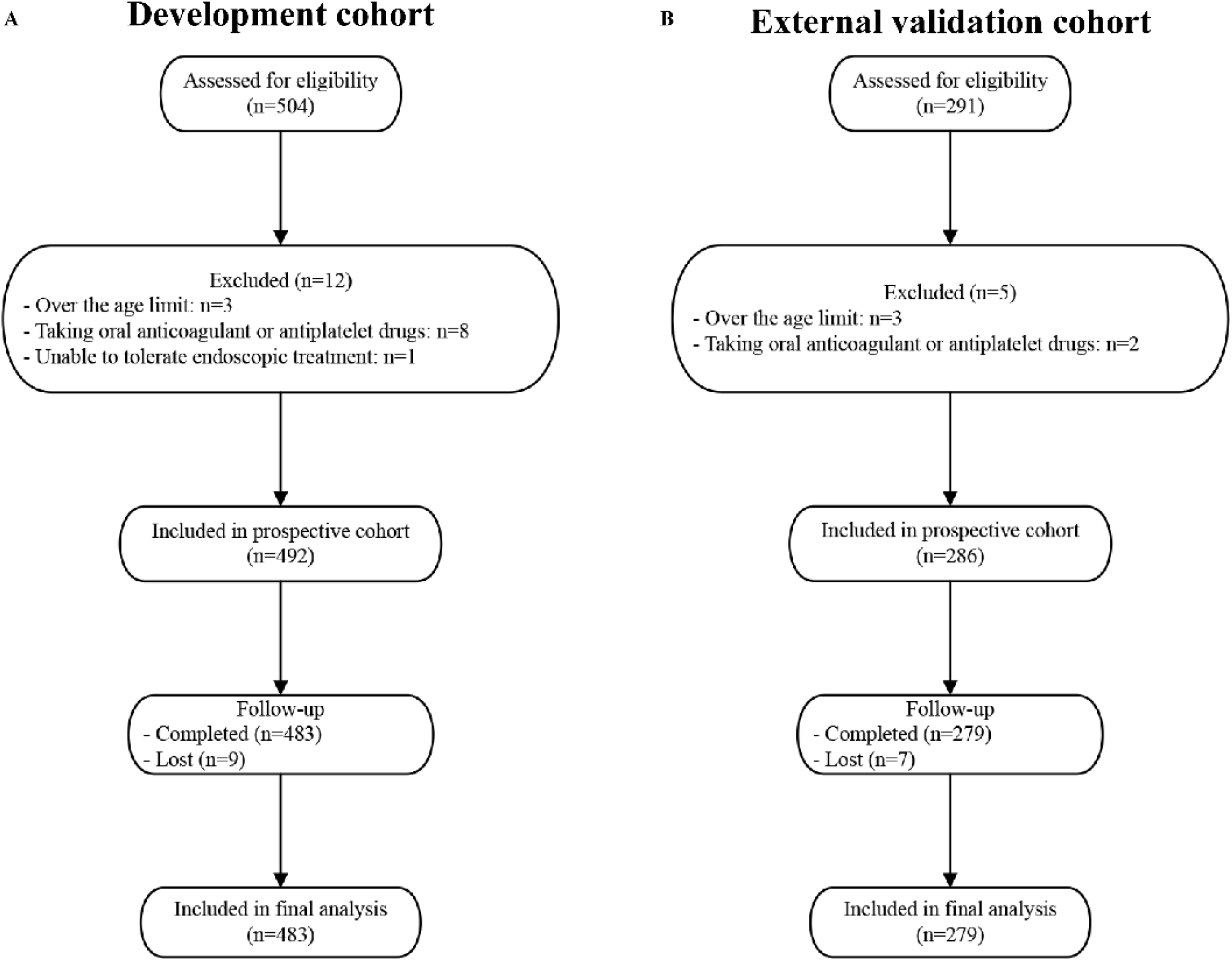
Study cohort flowchart. A. Development cohort. B. External validation cohort.

Statistical significance was defined as P < 0.05.

## RESULTS

### Patient characteristics and follow-up

A total of 504 patients with grade I internal hemorrhoids were prospectively enrolled in the development cohort. Twelve were excluded because of protocol-defined ineligibility, current anticoagulant or antiplatelet therapy, or inadequate retroflexed visualization. Nine were lost before completion of the minimum follow-up. Consequently, 483 patients were included in the primary development analysis (Figure 1A, Supplementary Table S9).

The cohort comprised 50.9% (246/483) male patients, with a median age of 52.5 years (interquartile range, 11.8 years). During the full available follow-up, recurrence was documented in 23.8% (115/483). Age did not differ between the recurrence and non-recurrence groups (55.0 [interquartile range, 21.0] vs 55.0 [interquartile range, 18.0] years, *P* = 0.82). The proportion of male patients was higher in the recurrence group (61.7% [71/115]) than in the non-recurrence group (47.6% [175/368], *P* = 0.008). Observed follow-up was shorter in patients with recurrence because observation ended at relapse (27.4 ± 9.3 vs 61.0 ± 11.0 months, P < 0.0001) (Table 1).

**Table 1.** Baseline clinical characteristics of patients in recurrence and no recurrence groups.

| Variable | Recurrence (*n* = 115) | No recurrence (*n* = 368) | *P* |
| --- | --- | --- | --- |
| Age, years, median (IQR) | 55.0 (21.0) | 55.0 (18.0) | 0.82 |
| Male sex, *n* (%) | 71 (61.7) | 175 (47.6) | <sup>b</sup>0.008 |
| Observed follow-up, months (mean ± SD) | 27.4 ± 9.3 | 61.0 ± 11.0 | <sup>d</sup><0.0001 |

Age is presented as median (interquartile range); observed follow-up is presented as mean ± SD. Median potential follow-up for the entire cohort (reverse Kaplan-Meier): 61.0 months (95%CI: 58.9–63.1).<sup>a</sup>*P* < 0.05, <sup>b</sup>*P* < 0.01, <sup>c</sup>*P* < 0.001, <sup>d</sup>*P* < 0.0001.

The reverse Kaplan-Meier median potential follow-up was 61.0 months (95%CI: 58.9–63.1). Of 483 patients, 368 were censored without a documented recurrence. Censoring occurred predominantly after prolonged observation (Supplementary Figure S2). Ninety-seven recurrences occurred by the prespecified 36-month prediction horizon and 18 additional recurrences occurred thereafter, explaining the total of 115 events. The administrative censoring date was December 31, 2025. The exact maximum period follow-up was 87 months.

Kaplan-Meier estimates showed cumulative recurrence rates of 0.6% (3/483), 2.5% (12/483), 10.1% (49/483), and 20.0% (97/483) at 6, 12, 24, and 36 months, respectively. No severe procedure-related adverse events were observed during treatment or follow-up.

### Endoscopic characteristics

At baseline, the recurrence group had more hemorrhoids (P < 0.0001); 86.1% (99/115) had three or more hemorrhoids compared with 52.7% (194/368) in the non-recurrence group. The recurrence group also had larger maximum hemorrhoid diameters (P < 0.0001) and more grade 2–3 red color signs (38.3% vs 17.1%, P < 0.0001). Mucosal erosion (*P* = 0.25) and dentate-line injury (*P* = 0.26) did not differ significantly (Table 2).

**Table 2.**
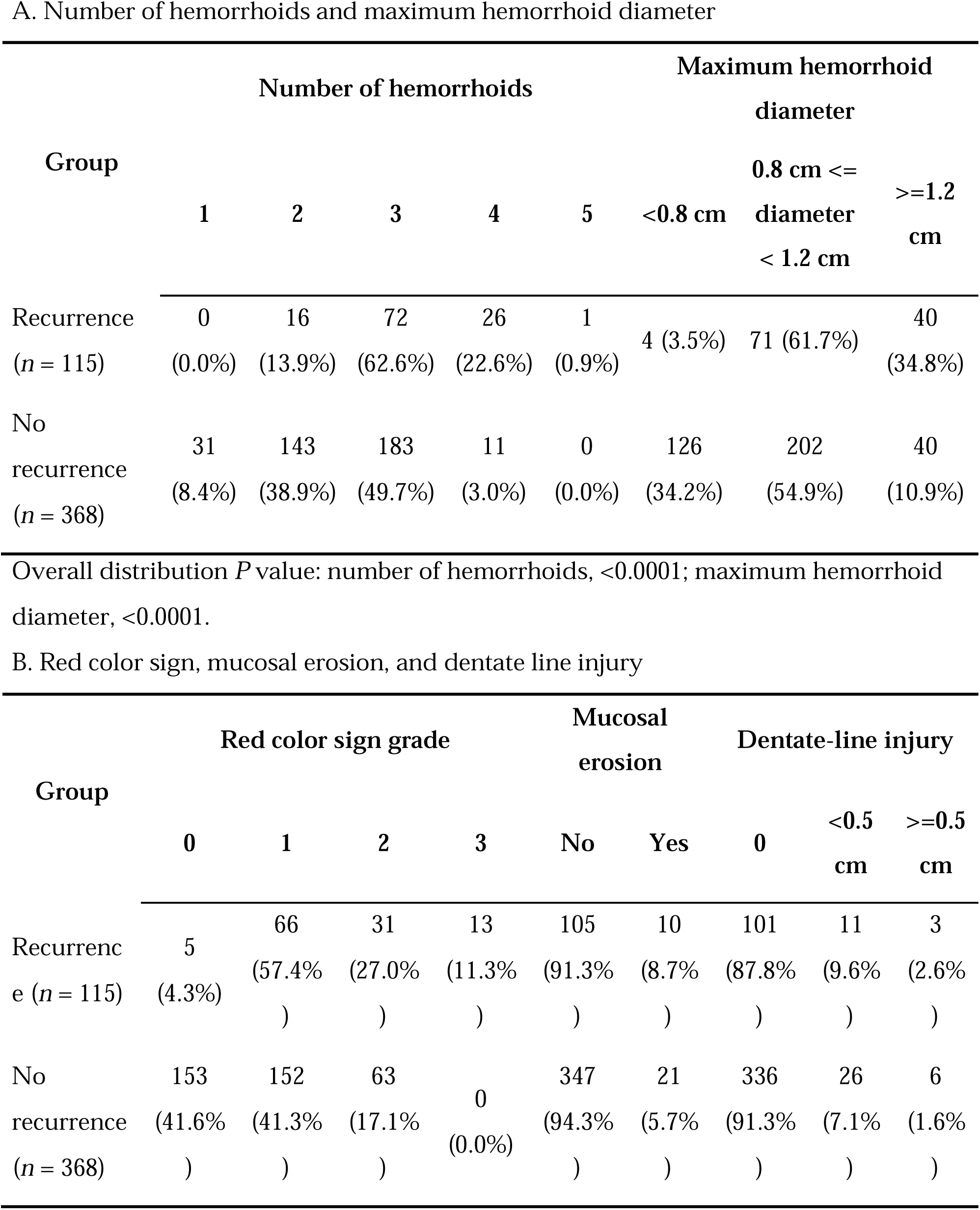
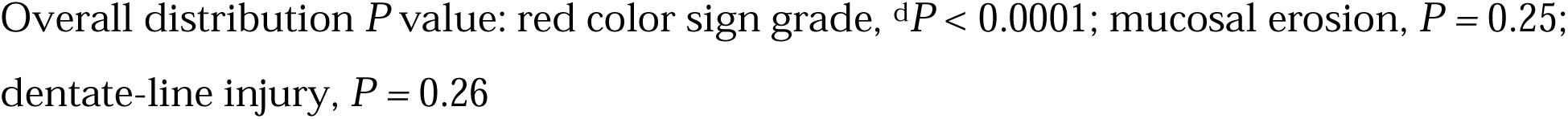
Endoscopic characteristics of patients in recurrence and no recurrence groups.

### Endo-HRS model construction

Number of hemorrhoids, maximum hemorrhoid diameter, and red color sign grade were strong predictors in the multivariable model (all P < 0.0001). Male sex did not reach conventional statistical significance (*P* = 0.08) and is therefore interpreted as a borderline, exploratory component rather than an established independent recurrence factor. It was retained because it was prespecified, its effect direction was clinically plausible, men were overrepresented among recurrent cases, and inclusion did not materially impair overall model performance. All four variables satisfied the proportional hazards assumption (Figure 2).

**Figure 2.**
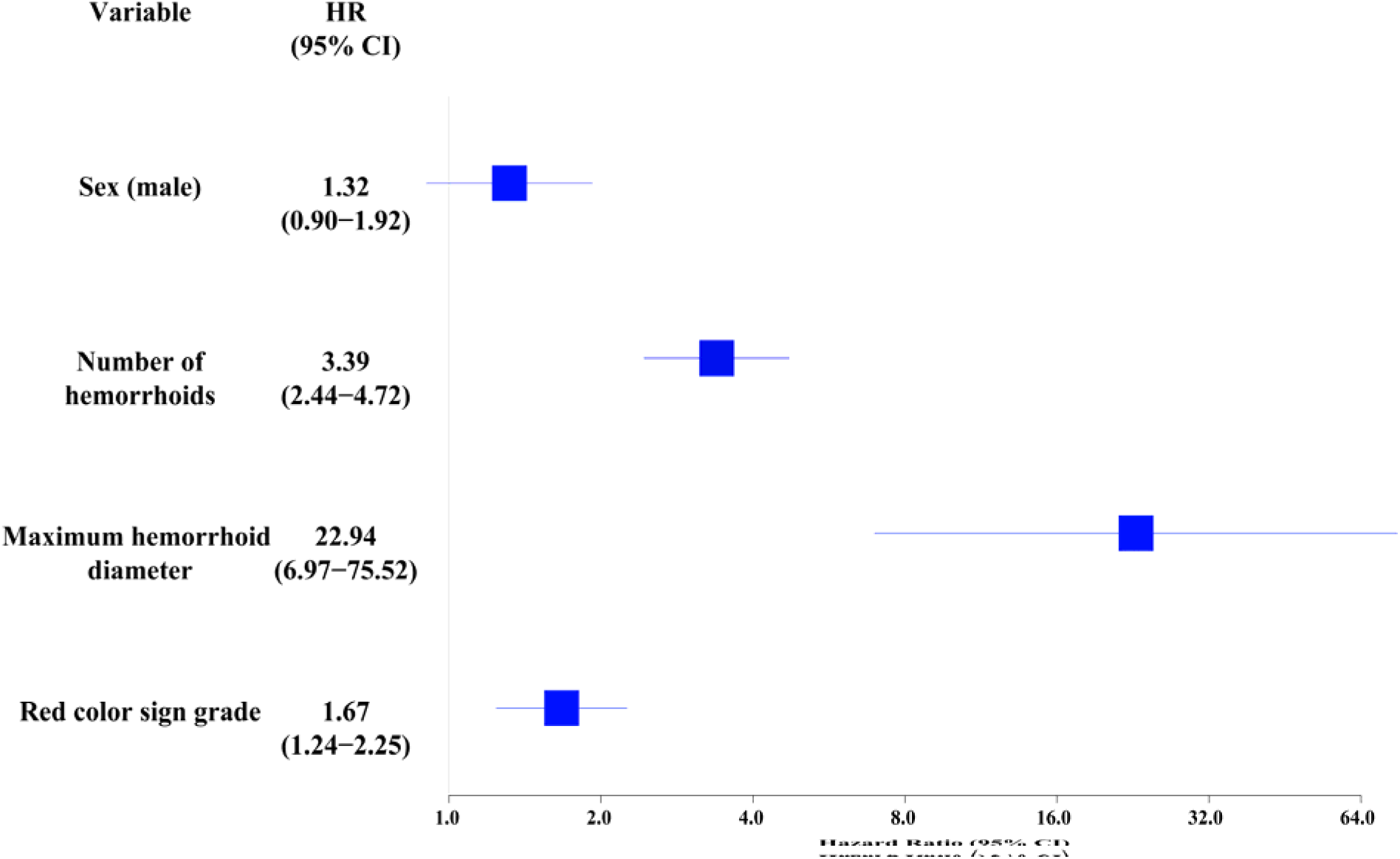
Forest plot of variables in the prediction model.

The continuous Cox model had an apparent C-index of 0.82 (SE 0.017), with time-dependent AUCs of 0.92, 0.93, and 0.88 at 12, 24, and 36 months, respectively (Supplementary Figure S3 and Supplementary Table S5). The 36-month AUC of 0.86 refers to the simplified three-tier clinical classification rather than the continuous Cox model. At 36 months, the reported calibration-in-the-large was -2.51 and the calibration slope was 1.34, indicating systematic underestimation of absolute risk; the 36-month Brier score was 0.112. Bootstrap optimism-corrected performance and external metrics are summarized in Supplementary Table S5. Decision curve analysis showed a positive net benefit across threshold probabilities of 2-50%, consistently outperforming both the treat-all and treat-none strategies (Supplementary Figure S4).

The Cox coefficients were rescaled relative to the coefficient for sex and rounded to generate the clinical weights. Because diameter is entered in centimeters as a continuous value, Endo-HRS (Formula 1) is a weighted continuous score and may contain decimals. The complete model specification and the absolute-risk equation are provided in Supplementary Table S4.

Endo-HRS = 1 × sex (male) + 4 × number of hemorrhoids + 11 × maximum hemorrhoid

diameter (cm) + 2 × red color sign grade (0–3)

Formula 1. Endo-HRS weighted continuous score.

Quartile-based groups were explored initially (Supplementary Table S10). For clinical presentation, groups A (< 18.2), B (18.2 to < 27.4), and C (≥ 27.4) were defined by lower rule-out and upper rule-in thresholds (Supplementary Table S11). The Youden-optimized threshold of 22.8 is a separate binary reference. The three-tier classification had a 36-month AUC of 0.86 (Figure 3A), recurrence-free interval decreased across groups (Figure 3B), and a nomogram estimated individual 36-month recurrence probability (Figure 3C).

**Figure 3.**
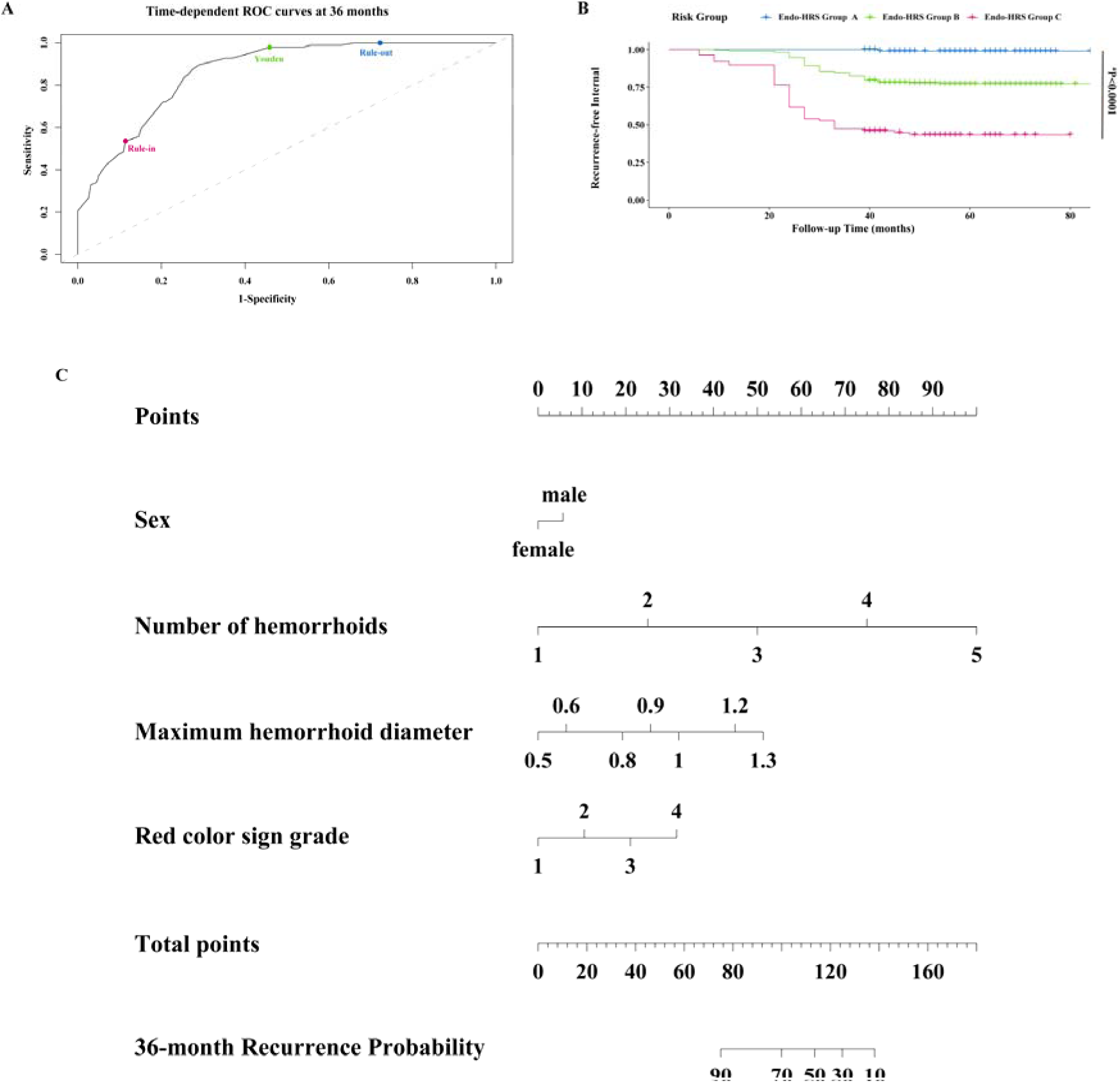
Model performance and clinical application of Endo-HRS. A: Time-dependent receiver operating characteristic curve for 36-month recurrence. The bronze, green, and blue points represent the rule-in, Youden-optimized, and rule-out thresholds, respectively. The Youden threshold (22.8) is a binary reference, whereas the lower and upper thresholds define the three-tier groups. B: Kaplan-Meier recurrence-free interval curves for groups A, B, and C. C: Nomogram estimating individual 36-month recurrence probability. AUC: Area under the curve.

The web calculator outputs the total score and risk group (Supplementary Figure S5). A printable calculation sheet with a worked example is provided in Supplementary Table S6, allowing manual use if the website is temporarily unavailable. Long-term maintenance and offline access are described in the Methods.

### External validation and exploratory biological verification

In the external cohort, 291 patients were screened and 279 were analyzed after five exclusions and seven losses before completion of the minimum follow-up (Figure 1B). The cohort was recruited at five contributing external centers and included 60 recurrence events during a median potential follow-up of 33.2 months (95% CI 32.4-34.1). Endo-HRS showed a bootstrap-corrected C-index of 0.900 (95%CI: 0.862–0.933), with time-dependent AUCs of 0.90, 0.94, and 0.95 at 24, 36, and 48 months, respectively (Figure 4A). Calibration was visually acceptable, with the slope of 0.97, and 36-month Brier score of 0.10 (Supplementary Figure S6 and Table S5). Group C had a shorter recurrence-free interval than groups A and B (P < 0.0001; Figure 4B).

**Figure 4.**
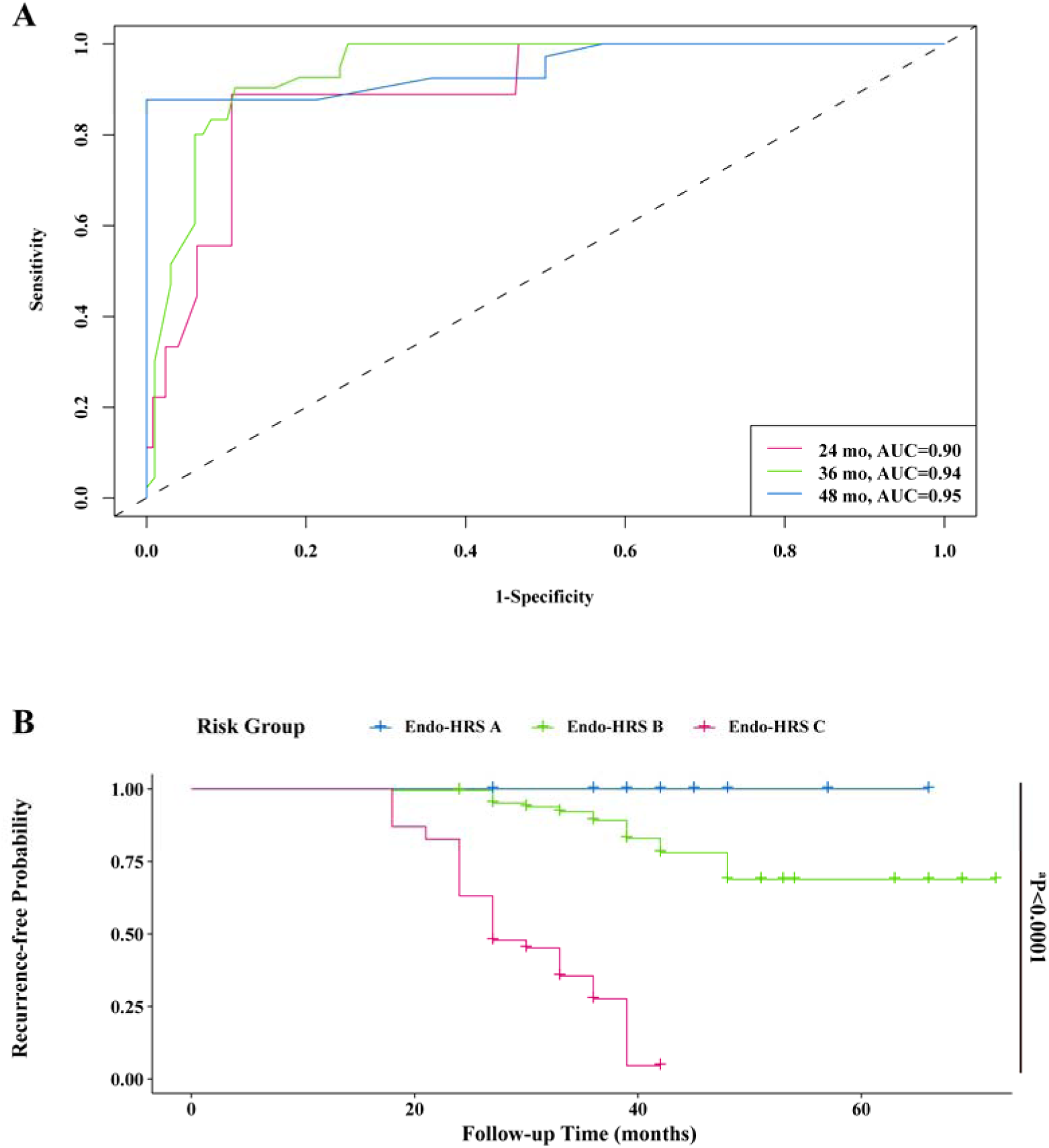
External validation of the Endo-HRS model. A. Time-dependent receiver operating characteristic (ROC) curves for recurrence prediction at 24 (bronze), 36 (green), and 48 (blue) months. B. Kaplan-Meier recurrence-free interval curves for Endo-HRS groups A (blue), B (green), and C (bronze).

In the sensitivity analysis restricted to recurrent bleeding with endoscopically confirmed hemorrhoids, predictor effects and model performance remained directionally consistent and verified C-index, AUC, and calibration values remained stable (Supplementary Table S12).

The exploratory biological-verification cohort included 40 participants: 9 in group A, 24 in group B, and 7 in group C (Supplementary Table S13). All contributed endoscopic images; quantitative Masson analysis included 15 adequate specimens, with 5 per group. The small and imbalanced group sizes limit statistical power.

Endoscopic images in higher-risk groups showed more prominent red color signs, more hemorrhoids, and larger diameters (Figure 5A–C). These cross-sectional findings were consistent with the variables encoded in Endo-HRS but do not constitute independent validation of predictive performance.

**Figure 5.**
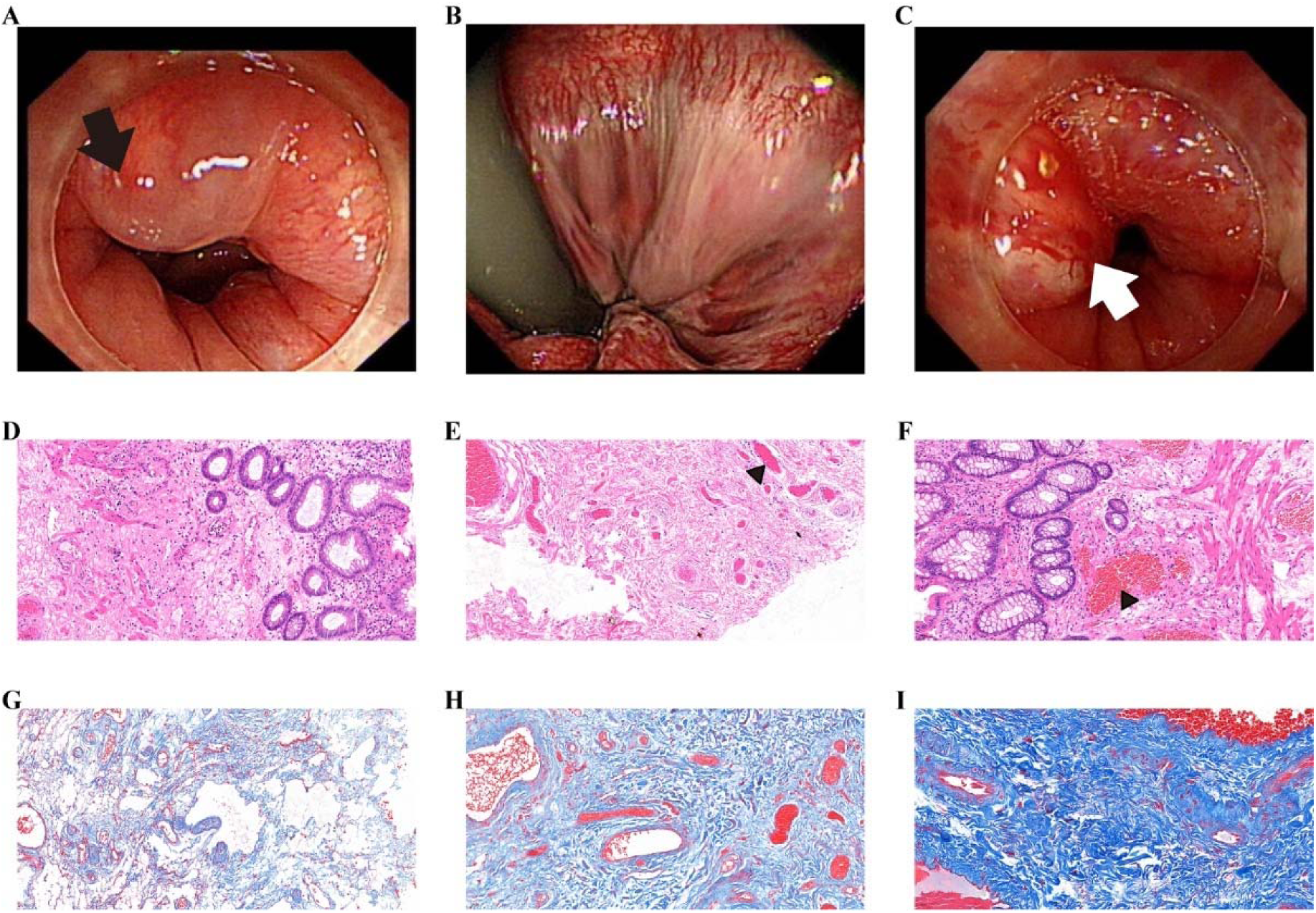
Exploratory multimodal biological verification of Endo-HRS. A–C: Endoscopic images from groups A, B, and C, respectively (black arrows: hemorrhoids; white arrows: red color signs). D–F: Representative hematoxylin-eosin sections (20×; scale bar = 50 μm; black triangles: enlarged vessels). G–I: Representative Masson’s trichrome sections (20×; scale bar = 50 μm). These images provide exploratory biological context and do not constitute independent predictive validation.

Representative histopathology showed greater dilation and congestion of submucosal venules in group C (Figure 5D–F). Masson’s trichrome staining showed higher collagen-proportionate area in higher-risk groups; group C values were 14.6-fold and 2.6-fold those of groups A and B, respectively (Figure 5G-I, Supplementary Table S14 and Figure S7). These exploratory, cross-sectional observations support biological plausibility but do not establish that vascular or extracellular-matrix changes cause recurrence.

## DISCUSSION

This prospective multicenter study evaluated long-term recurrence after 1% polidocanol foam sclerotherapy for grade I internal hemorrhoids and developed a weighted continuous score from standardized endoscopic features. Building on evidence supporting endoscopic polidocanol therapy[12–14, 31], the study addresses the unmet need for transparent recurrence-risk estimation while providing external evaluation and exploratory biological support.

Recurrence increased progressively over time, with cumulative rates of 2.5%, 10.1%, and 20.0% at 12, 24, and 36 months, respectively. These findings are broadly consistent with prior reports[6, 32], although previous studies largely used 3% polidocanol and focused on grade II-III disease[6, 32–35]. In our cohort, 1% polidocanol foam maintained durable efficacy in grade I disease, and no severe procedure-related adverse events were recorded.

Despite the substantial burden of hemorrhoidal disease[36], validated tools for individualized prediction of recurrence after endoscopic therapy remain scarce. The Goligher classification is based primarily on the degree of prolapse and has shown only fair interobserver agreement; moreover, it does not capture other potentially relevant phenotypic dimensions, including bleeding, hemorrhoid number and size, and mucosal surface changes[17]. Alternative multidimensional and patient-reported frameworks broaden the assessment of anatomy, bleeding, symptom burden, and health-related quality of life, but they were developed mainly for disease characterization, severity assessment, or treatment selection rather than individualized recurrence prediction[18–21]. Endo-HRS was designed to address this gap by combining sex with three standardized endoscopic features—number of hemorrhoids, maximum hemorrhoid diameter, and red color sign grade—which represent complementary dimensions of disease extent, lesion size, and mucosal surface appearance. The selection of these variables is supported by previous endoscopic phenotyping frameworks: colonoscopic assessments based on lesion extent, form or size, and red color signs have demonstrated associations with clinical symptoms, particularly bleeding[22]. Contemporary PBR and LDRF frameworks similarly emphasize hemorrhoid diameter and mucosal surface risk features, while recommending systematic documentation of hemorrhoid size, number, distribution, and surface abnormalities[23]. Standardized assessment using both antegrade and retroflexed views, together with an endoscopic scale marker for size estimation, was intended to reduce measurement ambiguity and improve operational consistency. Endo-HRS should therefore be regarded as complementary to, rather than a replacement for, established prolapse- and symptom-based systems. Nevertheless, formal assessment of inter- and intraobserver agreement remains necessary, and the discrimination, calibration, and clinical utility of the Endo-HRS model should be confirmed in prospective external cohorts.

Male sex was retained as a borderline exploratory predictor rather than being interpreted as a proven independent determinant of post-sclerotherapy recurrence. Direct evidence specifically linking male sex to recurrence after 1% polidocanol foam sclerotherapy for grade I internal hemorrhoids remains limited. However, in the prospective multicenter SCLEROFOAM study of 3% polidocanol foam for second-degree hemorrhoidal disease, gender was associated with recurrence, with most recurrent cases occurring in men[32]. In the 3-year follow-up of the same multicenter cohort, gender remained associated with recurrence, and recurrent events were more frequent in men than in women[35]. Broader evidence from a systematic review and the international CHORUS real-world study also identified male gender among recurrence-related or severity-related factors in hemorrhoidal disease[20, 37]. In addition, genome-wide analyses suggest that hemorrhoidal disease has a genetic component involving vascular, smooth-muscle, epithelial/endothelial, and extracellular-matrix pathways, supporting biological heterogeneity rather than a purely mechanical phenotype[38]. Therefore, male sex was retained in Endo-HRS as a prespecified, borderline exploratory component, not as an established independent recurrence determinant. Its contribution should be re-estimated during future model updating in larger, diverse cohorts, and the performance of models with and without sex should be compared.

The lower and upper operating thresholds provide a simple three-tier presentation, while 22.8 remains a separate Youden-optimized binary reference. These groups may aid communication of relative risk and hypothesis generation for future management studies. They should not yet be interpreted as prospectively validated instructions for surveillance intensity or retreatment, because no impact trial compared Endo-HRS-guided care with usual practice.

The exploratory multimodal component provides biological context rather than definitive validation. Hemorrhoidal disease is increasingly recognized as a degenerative disorder of the anal cushion characterized by vascular remodeling and extracellular-matrix dysregulation[18, 19, 39]. Chronic venous hypertension and microcirculatory disturbance may lead to persistent venous dilation and structural remodeling of hemorrhoidal plexuses. At the molecular level, imbalance in extracellular-matrix turnover, including dysregulated collagen synthesis and degradation mediated by matrix metalloproteinase pathways, may contribute to tissue weakening and remodeling[39]. Genome-wide analyses further support a genetically determined component involving vascular development, smooth-muscle function, epithelial/endothelial biology, and extracellular-matrix organization pathways[38]. In addition, extracellular-matrix dynamics and collagen remodeling are central regulators of connective-tissue structure and vascular homeostasis[40], while collagen fiber reorganization, mechanical stress propagation, and cell-matrix interactions can drive tissue remodeling and structural adaptation[40, 41]. Within this framework, the observed venous congestion, hemorrhoid burden, red color signs, and collagen differences likely represent macroscopic manifestations of underlying vascular and extracellular-matrix remodeling. However, the cross-sectional nature of histological assessment precludes inference on temporal or causal relationships between collagen deposition and recurrence. These findings are therefore hypothesis-generating and require validation in longitudinal and mechanistic studies.

This study has several limitations. First, development was restricted to Chinese patients with grade I internal hemorrhoids treated with 1% polidocanol foam, limiting generalizability to other populations, grades, and therapies. Second, constipation, occupational exposure, diet, pregnancy and childbirth history, and other patient-level factors were not prospectively captured with standardized definitions; future model updating should evaluate their incremental value. Third, the high-specificity endoscopic recurrence definition reduces misclassification from non-hemorrhoidal symptoms but may underestimate the broader symptomatic burden. Conceptual overlap remains possible between baseline red color sign and follow-up vascular abnormalities, despite temporal separation, blinded adjudication, and the prespecified sensitivity analysis. Fourth, endoscopic variables were independently evaluated during routine clinical assessment, with discrepancies resolved by consensus. However, only the consensus findings were retained in the final analytical dataset, and the individual pre-consensus ratings were not archived. Consequently, formal measures of interobserver reproducibility (e.g., intraclass correlation coefficients or kappa statistics) could not be calculated retrospectively. Although standardized training, prespecified definitions, antegrade and retroflexed views, scale-marker-assisted measurement, and consensus review were used to reduce assessment variability, future prospective studies should retain independent readings and report interobserver agreement for these endoscopic predictors. Fifth, the use of univariable screening and backward selection may have produced unstable or optimistic coefficients; although bootstrap validation was performed, residual optimism cannot be fully excluded. Finally, the exploratory biological cohort was small and imbalanced, precluding causal inference. The web calculator is supplementary to the printed formula and requires ongoing maintenance.

## CONCLUSION

Endo-HRS is a weighted continuous endoscopy-based score with encouraging development and external-validation performance after foam sclerotherapy for grade I internal hemorrhoids. It may aid recurrence-risk stratification and support the design of future risk-adapted follow-up studies; prospective impact evaluation is required before it is used to direct surveillance or retreatment.

## Data Availability

All data produced in the present study are available upon reasonable request to the authors

## ACKNOWLEDGEMENTS

The authors thank all patients for participating and for permitting the use of their treatment and follow-up data. The authors also thank the medical statistics team of Xinhua Hospital for methodological support. All authors take responsibility for the integrity of the work.

## Author contributions

Zhang FY and Xu LM contributed equally to this work; Zhang FY, Xu LM, Hu Y and Shen F conceived and designed the study; Gao FY, Wang W, Lin WL, Zhang H, Wang QJ, Jin AJ, Yang RP, Qu CY, Zhang Y, Li ZH, Wang D, Shi CC, Shen TB and Shen F collected clinical, endoscopic and follow-up data; Wang KZ performed the pathological assessment; Zhang FY and Xu LM analyzed the data and drafted the manuscript; Hu Y and Shen F jointly supervised the study and critically revised the manuscript; all authors interpreted the data, approved the final manuscript, and agree to be accountable for all aspects of the work.

## Supported by

the Discipline Peak-Climbing Plan of Xinhua Hospital Affiliated to Shanghai Jiao Tong University School of Medicine, No. XKPF2024B404; the Shanghai Science and Technology Commission Innovation Ecosystem Construction Plan ’Domestic Science and Technology Cooperation’ Project, No. 25010702300; and the National Natural Science Foundation of China, No. 82472899.

## Clinical trial registration statement

This study was prospectively registered at ClinicalTrials.gov (NCT04398823).

## Biostatistics statement

The statistical methods of this study were reviewed by biomedical statisticians from the Clinical Research Unit of Xinhua Hospital.

### STROBE statement

The authors have read the STROBE Statement—checklist of items, and the manuscript was prepared and revised according to the STROBE Statement—checklist of items.

## Footnotes

### Institutional review board statement

The study was reviewed and approved by the Ethics Committee of Xinhua Hospital Affiliated to Shanghai Jiao Tong University School of Medicine (Approval No. XHEC-C-2020-003-1) and by the ethics committees of the participating centers.

### Informed consent statement

All study participants or their legal guardians provided informed written consent prior to study enrollment. Due to the large sample size, only a representative de-identified informed consent form is provided. All original signed informed consent forms are securely archived at our institution and are available upon request.

### Preprint statement

A preprint version of this study has been made publicly available in medRxiv (https://doi.org/10.1101/2025.10.28.25338987).

### Data sharing statement

The deidentified data that support the findings of this study are available from the corresponding authors upon reasonable request, subject to institutional and ethical approval.

### STROBE statement

The authors have read the STROBE Statement-checklist of items, and the manuscript was prepared and revised according to the STROBE Statement-checklist of items.

### AI-assisted technologies statement

ChatGPT (OpenAI; accessed June 2026) was used solely for linguistic refinement and formatting assistance. No artificial intelligence tool was used to generate research data, perform statistical analyses, interpret results, or formulate conclusions. All AI-assisted text was critically reviewed and revised by the authors.

### Open-Access

This article is an open access article distributed under the terms and conditions of the Creative Commons Attribution-NonCommercial (CC BY-NC 4.0) license. No commercial re-use.

### Corresponding Author’s Membership in Professional Societies

Ye Hu, Member of the Shanghai Anti-Cancer Association; Member of the Small Bowel Study Group, Digestive Endoscopy Branch, Shanghai Medical Association Feng Shen, Vice Chairperson of the Young Committee of the Digestive Endoscopy Branch of the Shanghai Medical Association, Member of the Hepatitis Disease Public Education Committee of the Chinese Medical Association, Young Member of the Internal Medicine Branch of the Shanghai Medical Association.

### Specialty type

Gastroenterology and hepatology.

### Country/Territory of origin

China.

