## Supplementary material for "Endoscopic score predicting recurrence after foam sclerotherapy in grade I internal hemorrhoids: Development and external validation": SI

**SUPPLEMENTARY MATERIALS**

**SUPPORTING INFORMATION LEGENDS**

**Figure S1. Definition and representative endoscopic images of the “red color sign”.** The “red color sign” refers to endoscopic vascular abnormalities characterized by dilated, tortuous, and irregularly distributed capillaries on the surface of internal hemorrhoids. It was graded on a scale of 0 to 3 based on severity: grade 0 (absent), grade 1 (scattered red dots), grade 2 (linear red streaks), and grade 3 (confluent areas of red congestion).

Panel A: Grade 0 (absent).

Panel B: Grade 1 (black arrows indicate red dots).

Panel C: Grade 2 (white arrows indicate linear streaks).

Panel D: Grade 3 (arrow in a black-bordered box indicates an area of confluent congestion).

**Figure S2. Follow-up duration among censored patients.** Histogram showing the distribution of time to censoring, which includes both administrative censoring at the study end and loss to follow-up.

**Figure S3. Time-dependent receiver operating characteristic (ROC) curves for recurrence prediction at 12, 24, and 36 months, with corresponding area under the curve (AUC) values of 0.92, 0.93, and 0.88, respectively.** Abbreviations: mo: months.

**Figure S4. Decision curve analysis (DCA) of development cohort and validate cohort.** A. DCA of development cohort. B. DCA of validate cohort.

**Figure S5. Interface of the Endo-HRS calculator.** The web-based calculator allows clinicians to input four parameters, including sex of patients, number of hemorrhoids, maximum hemorrhoid diameter (cm), and grade of red color sign. The Endo-HRS is automatically computed, and patients are stratified into Endo-HRS group A (low risk), B (moderate risk), and C (high risk) according to the prespecified cut-off values.

**Figure S6. Calibration curves for the 36-month recurrence prediction in the external validation cohort, plotted using the Logit-LOESS method.** The gray line represents the ideal reference (perfect prediction), while the orange line shows the observed model performance.

**Figure S7. The percentage of Masson-stained collagen area across different experimental groups (n=5 per group).**

**Table S1** **Inclusion criteria of the study.**

**Inclusion criteria**

1. Age between 18-70 years;

2. Diagnosed with grade I hemorrhagic internal hemorrhoids;

3. Failure of conservative treatment;

4. No previous history of endoscopic treatment for hemorrhoids.

**Table S2 Exclusion criteria of the study.**

**Exclusion criteria**

1. Over the age of 70 or under the age of 18;

2. Diagnosed with grade II, III or IV internal hemorrhoids and external hemorrhoids;

3. Severe cardio- or pulmonary insufficiency;

4. Diagnosis of colon polyps > 1cm in diameter or at > 3 sites; or the presence of malignancy;

5. Diagnosis of inflammatory bowel disease or other perianal diseases;

6. Diagnosis of autoimmune diseases;

7. History of internal hemorrhoids surgery;

8. Allergy to treatment-related drugs;

9. Anticoagulant/Antiplatelet use within the past 7 days.

Chronic constipation or straining, prolonged sitting or standing, alcohol and spicy-food exposure, and pregnancy or childbirth history were not collected prospectively.

**Table S3.** **Questionnaire assessment of follow-up.**

| **Questions** | **Answers** | |
| --- | --- | --- |
| **Another bleeding after treatment?** | No, | Yes, when? |
| **If bleeding, the frequency** |  | |
| **Any other accompanying symptoms?** | No. | Yes: |
| **Any treatment?** | - Adequate fluid or fiber intake | |
|  | - Medication□ | |
|  | - Lifestyle changes | |
|  | - Physiotherapy | |
|  | - Endoscopic therapy | |
|  | - Surgery | |
|  | - Others: | |

**Table S4.** **Metrics of Cox proportional hazards model.**

| **Variable** | **β** | **SE** | **HR** | **95%CI** | ***P*** |
| --- | --- | --- | --- | --- | --- |
| **sex** | 0.28 | 0.19 | 1.32 | 0.90-1.92 | 0.152 |
| **number of hemorrhoids** | 1.22 | 0.17 | 3.39 | 2.44-4.72 | ^d^<0.0001 |
| **maximum diameter of hemorrhoids** | 3.13 | 0.61 | 22.94 | 6.97-75.72 | ^d^<0.0001 |
| **grade of red color sign** | 0.51 | 0.15 | 1.67 | 1.24-2.25 | ^d^<0.0001 |

**Baseline survival at 36 months: 0.90.**

^a^*P* < 0.05, ^b^*P* < 0.01, ^c^*P* < 0.001, ^d^*P* < 0.0001.

**Table S5.** **Apparent, internal, and external performance metrics of the cohorts.**

| **Metric** | **Development** | **Internal**  **(Bootstrap corrected)** | **External** |
| --- | --- | --- | --- |
| **C-index** | 0.82 | 0.82 | 0.90 |
| **AUC 12 months** | 0.92 | - | - |
| **AUC 24 months** | 0.93 | - | 0.90 |
| **AUC 36 months** | 0.88 | - | 0.94 |
| **AUC 48 months** | - | - | 0.95 |
| **Calibration slope** | 1.34 | 0.97 | 0.97 |
| **Brier score 36 months** | 0.11 | - | 0.10 |
| **IBS** | 0.09 | - | 0.09 |

**Table S6. Paper worksheet of the offline calculator**

| **Endo-HRS Recurrence Risk Calculator** | | |
| --- | --- | --- |
| **Predictors** | **Coefficient** | **Value** |
| **Sex (Male = 1, Female = 0)** | 1 |  |
| **Number of Hemorrhoids** | 4 |  |
| **Maximum Hemorrhoid Diameter (cm)** | 11 |  |
| **Grade of Red Color Sign (0, 1, 2, 3)** | 2 |  |
| **Results** | | |
| **Endo-HRS Score** |  | |
| **Risk group classification** | | |
| **Endo-HRS Group A (Low risk)** | < 18.2 | |
| **Endo-HRS Group B (Moderate risk)** | 18.2-27.4 | |
| **Endo-HRS Group C (High risk)** | ≥ 27.4 | |

Table S7 Baseline clinical characteristics of patients in external cohort.

| **Variable** | **Recurrence (*n* = 60)** | **No recurrence (*n* = 219)** | ***P*** |
| --- | --- | --- | --- |
| Age, years, median (IQR) | 54.0 (21.3) | 56.0 (19.0) | 0.301 |
| Male sex, n (%) | 41 (68.3) | 117 (53.4) | 0.055 |
| Observed follow-up, months (mean ± SD) | 29.5 ± 7.3 | 34.2 ± 6.8 | ^d^<0.0001 |

Median potential follow-up for the entire cohort (reverse Kaplan-Meier): 33.2 months (32.4-34.1).

^a^*P* < 0.05, ^b^*P* < 0.01, ^c^*P* < 0.001, ^d^*P* < 0.0001.

**Table S8.** **Relevant R packages used in the study.**

| **R packages** | **Version** | **R packages** | **Version** | **R packages** | **Version** |
| --- | --- | --- | --- | --- | --- |
| survival | 3.8.6 | timeROC | 0.4 | rms | 8.1.1 |
| riskRegression | 2026.03.11 | pec | 2025.06.24 | mice | 3.19.0 |
| tableone | 0.13.2 | dplyr | 1.2.1 | ggplot2 | 4.0.3 |

**Table S9 Missingness counts of variables.**

| **Variables** | **Missing number** |
| --- | --- |
| **sex** | **0** |
| **age** | **0** |
| **hemorrhoid number** | **0** |
| **maximum hemorrhoid diameter (cm)** | **0** |
| **red color sign grade (0-3)** | **0** |
| **mucosa erosion** | **0** |
| **dentate-line injury** | **0** |
| **recurrence status** | **0** |
| **follow-up period** | **0** |

**Table S10 Number of patients (*n*) and scores distribution of the Endo-HRS model quartile method.**

| **Risk** | **Low** | **Moderate** | **High** | **Very High** |
| --- | --- | --- | --- | --- |
| **_a_*n*** | 136 | 128 | 104 | 115 |
| **Score** | 11.5-20.6 | 20.6-24.8 | 24.8-27.2 | ≥ 27.2 |
| **_b_C-index 0.81** | | | | |

**_a_*n*: Number of patients; _b_C-index: Concordance index.**

**Table S11 Number of patients (*n*) and scores distribution of the clinically practical Endo-HRS model.**

| **Class** | **A** | **B** | **C** |
| --- | --- | --- | --- |
| **_a_*n*** | 110 | 284 | 89 |
| **Score** | ＜18.2 | 18.2-27.4 | ≥ 27.4 |

**_a_*n*: Number of patients.**

**Table S12 Sensitivity analysis (A) and model performance (B) of the refitting multivariable Cox model after excluding baseline red color sign.**

**(A)**

| **Variable** | **Endo-HRS model HR (95% CI)** | **P value** | **Sensitivity model HR (95% CI)** | ***P*** |
| --- | --- | --- | --- | --- |
| **Sex** | 1.32 (0.90-1.92) | 0.155 | 1.38 (0.94-2.01) | 0.096 |
| **Number of hemorrhoids** | 3.39 (2.44-4.72) | < 0.0001 | 3.75 (2.72-5.16) | ^d^<0.0001 |
| **Maximum diameter of hemorrhoids** | 22.94 (6.97-75.52) | < 0.0001 | 55.97 (19.11-163.85) | ^d^<0.0001 |
| **Grade of red color sign** | 1.67 (1.24-2.25) | < 0.0001 | - | Excluded |

^a^*P* < 0.05, ^b^*P* < 0.01, ^c^*P* < 0.001, ^d^*P* < 0.0001.

**(B)**

|  | **Endo-HRS model** | **Sensitivity model** |
| --- | --- | --- |
| **Harrell’s C-index** | 0.82 | 0.81 |
| **36-month AUC** | 0.86 | 0.87 |
| **Calibration slope (36 months)** | 1.34 | 1.34 |

**Table S13 Risk factors of selected patients for multi-modal verification.**

| **Groups** | | **Endo-HRS A (n=9)** | **Endo-HRS B (n=24)** | **Endo-HRS C (n=7)** | ***P*** |
| --- | --- | --- | --- | --- | --- |
| **Scores** | | 16.1±1.2 | 23.1±2.4 | 29.6±1.8 | ^d^*P* < 0.0001 |
| **Status** | **Recurrence** | 0 | 6 | 2 | ^b^P=0.003 |
|  | **No recurrence** | 9 | 18 | 5 |  |
| **Sex** | **Male** | 5 | 10 | 6 | P=0.15 |
|  | **Female** | 4 | 14 | 1 |  |
| **Number of hemorrhoids** | **2** | 9 | 3 | 0 | ^d^*P* < 0.0001 |
|  | **3** | 0 | 19 | 5 |  |
|  | **4** | 0 | 2 | 2 |  |
| **Maximum hemorrhoid diameter** | **<0.8cm** | 7 | 8 | 0 | ^d^*P* < 0.0001 |
|  | **0.8cm≤Dia.<1.2cm** | 2 | 15 | 3 |  |
|  | **≥1.2cm** | 0 | 1 | 4 |  |
| **Grade of red color sign** | **0** | 7 | 1 | 1 | ^b^P=0.003 |
|  | **1** | 2 | 17 | 2 |  |
|  | **2** | 0 | 4 | 3 |  |
|  | **3** | 0 | 2 | 2 |  |

^a^*P* < 0.05, ^b^*P* < 0.01, ^c^*P* < 0.001, ^d^*P* < 0.0001.

**Table S14 The Percentage of Masson-positive staining area in different groups** (mean ± SD, *n*=5).

| **Endo-HRS Group** | A | B | C |
| --- | --- | --- | --- |
| **Area (%)** | 3.30±1.88 | 18.76±3.27 | 48.10±3.88 |

**Figure S1**

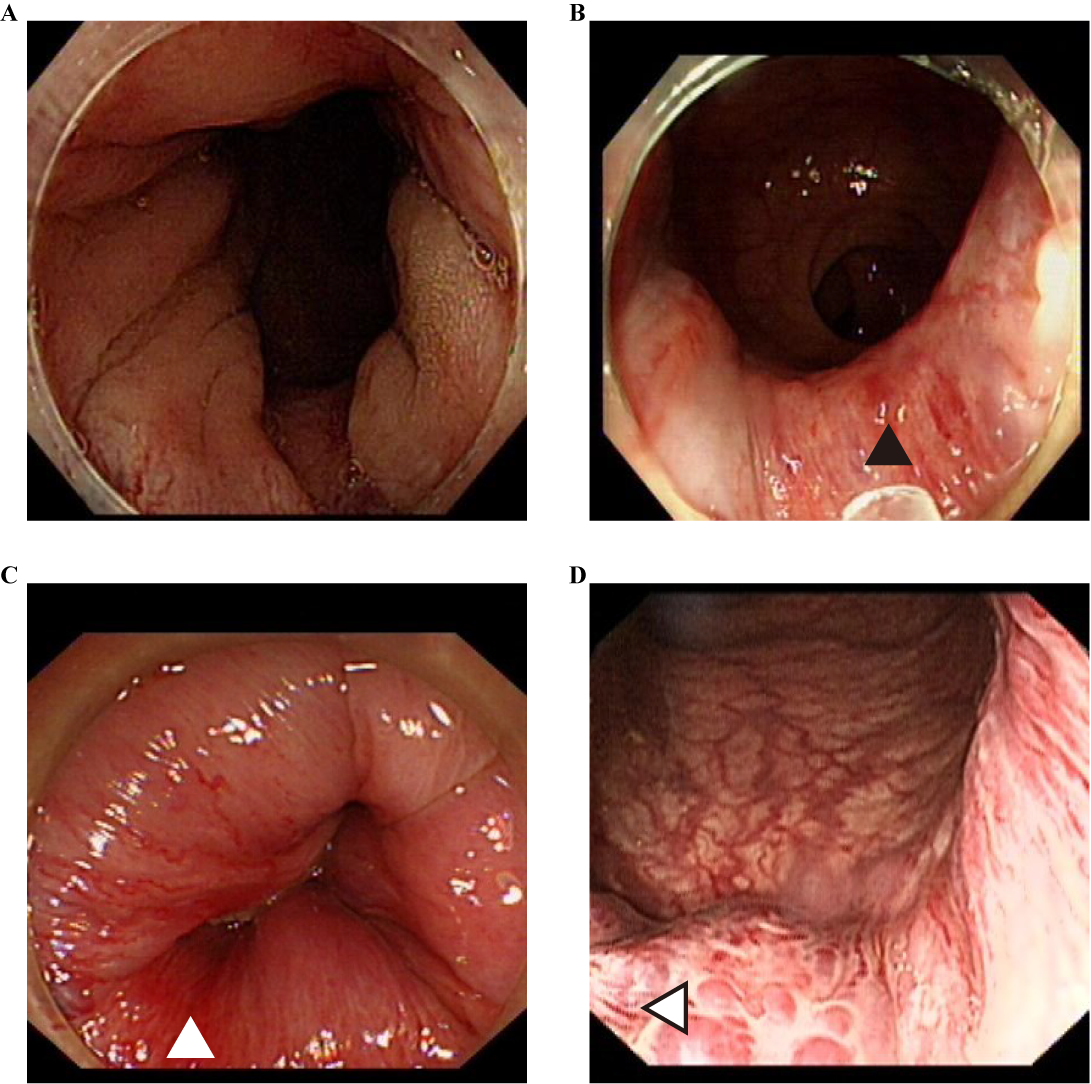

**Figure S2**

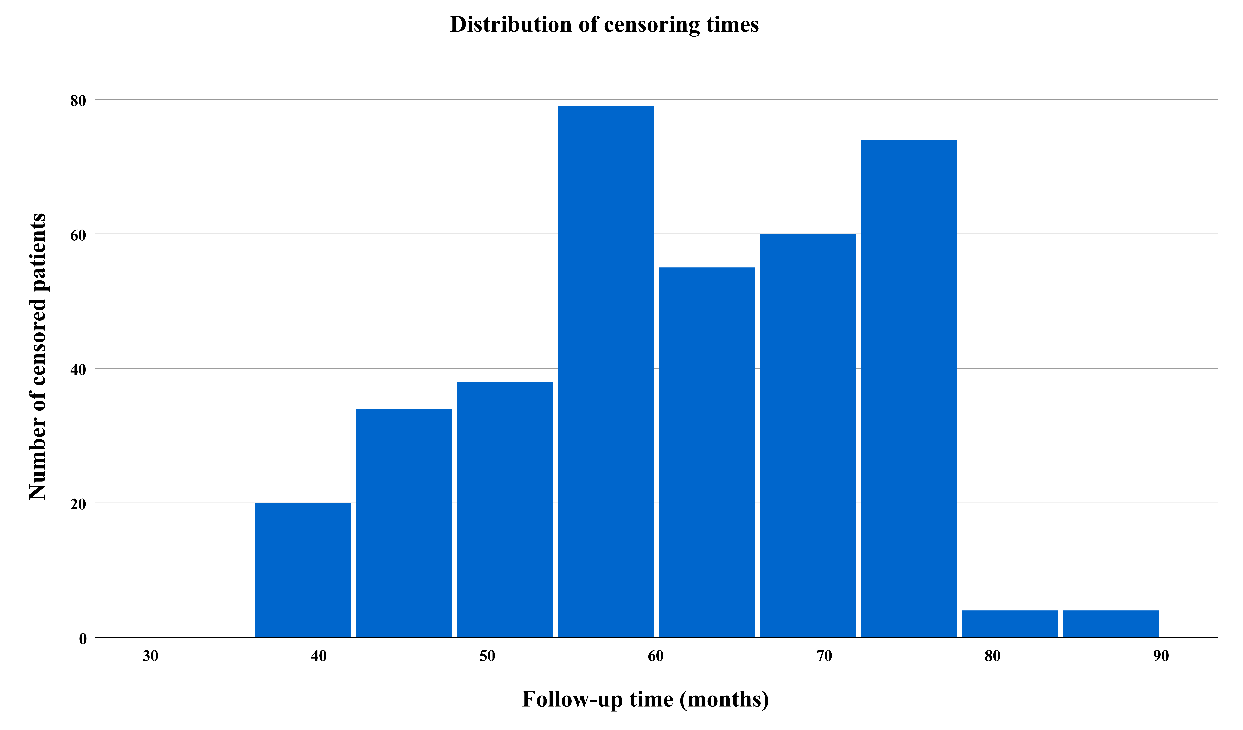

**Figure S3**

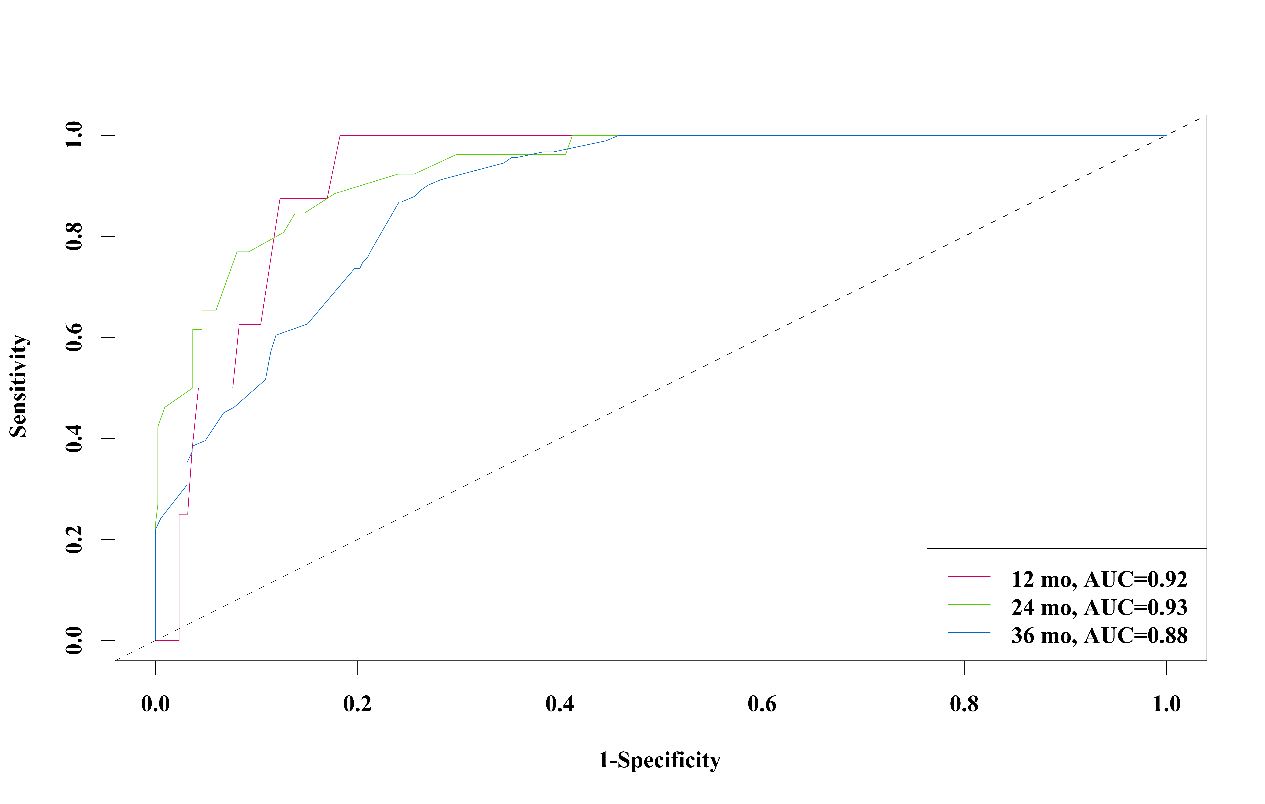

**Figure S4**

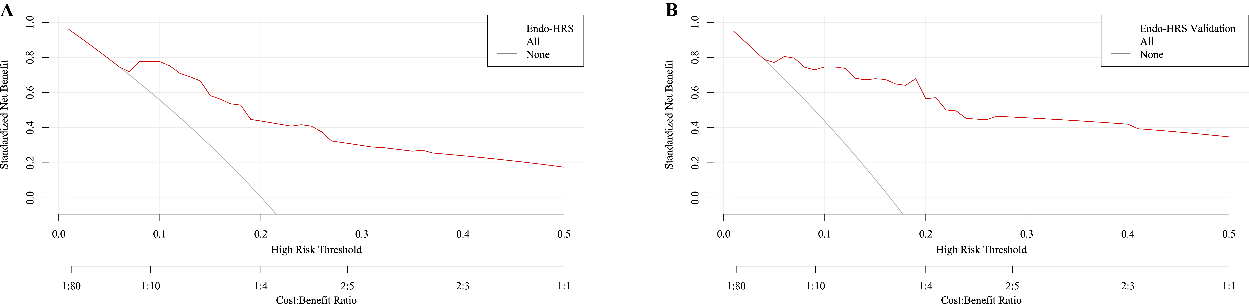

**Figure S5**

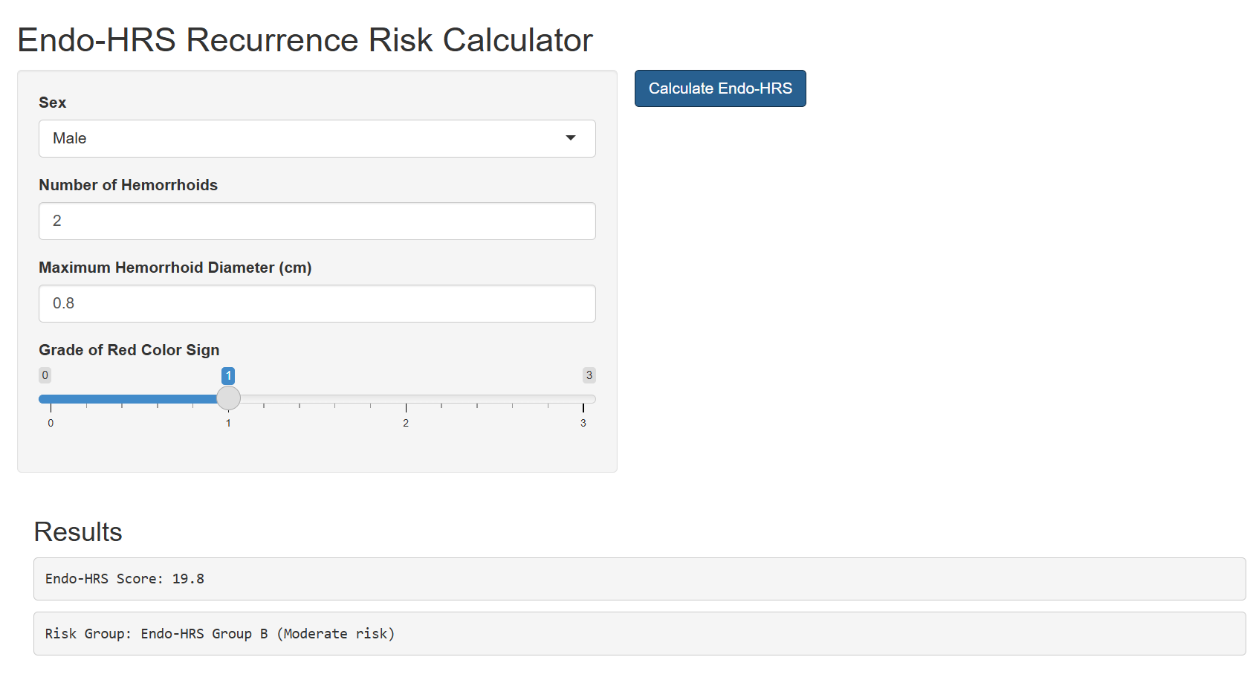

**Figure S6**

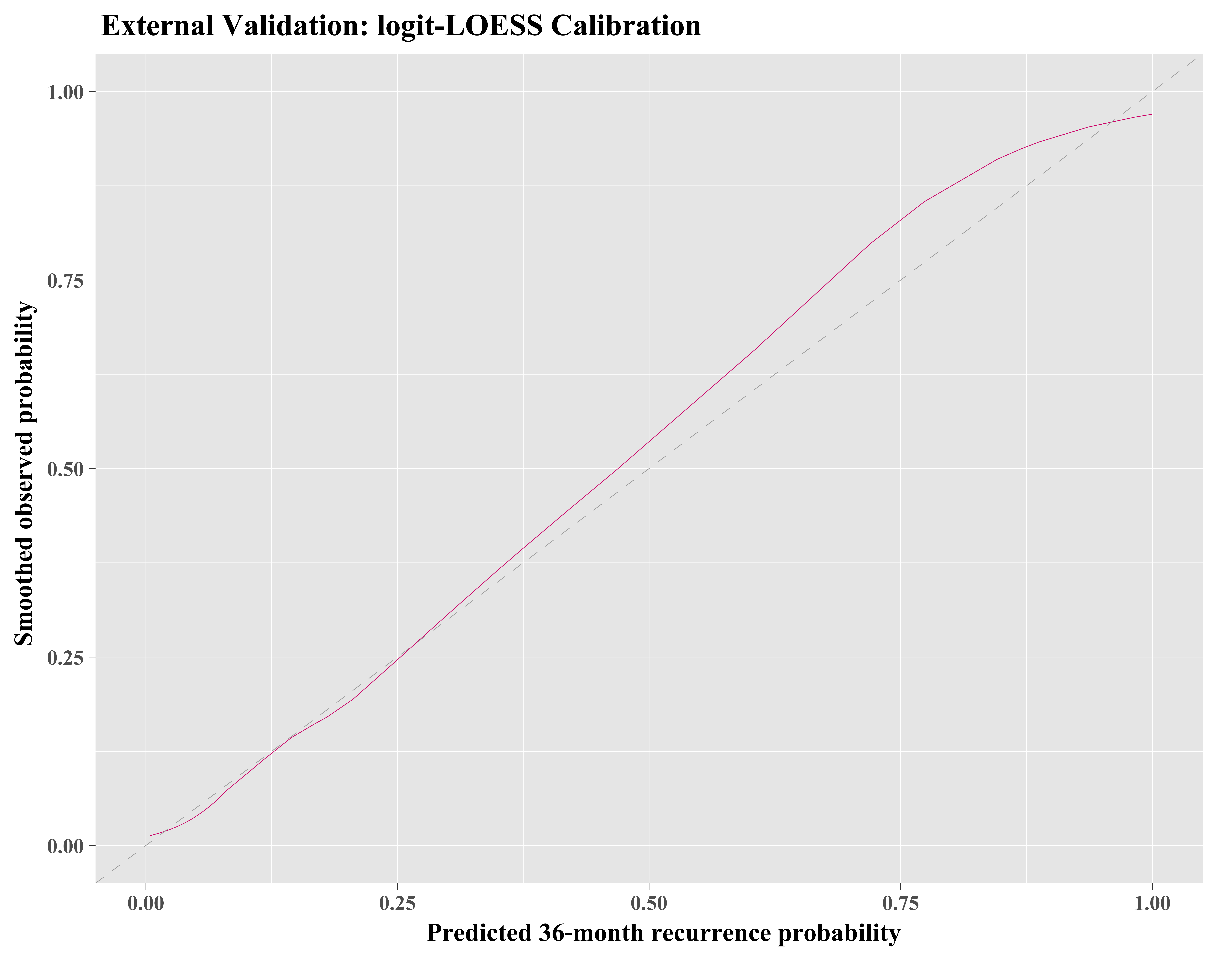

**Figure S7**

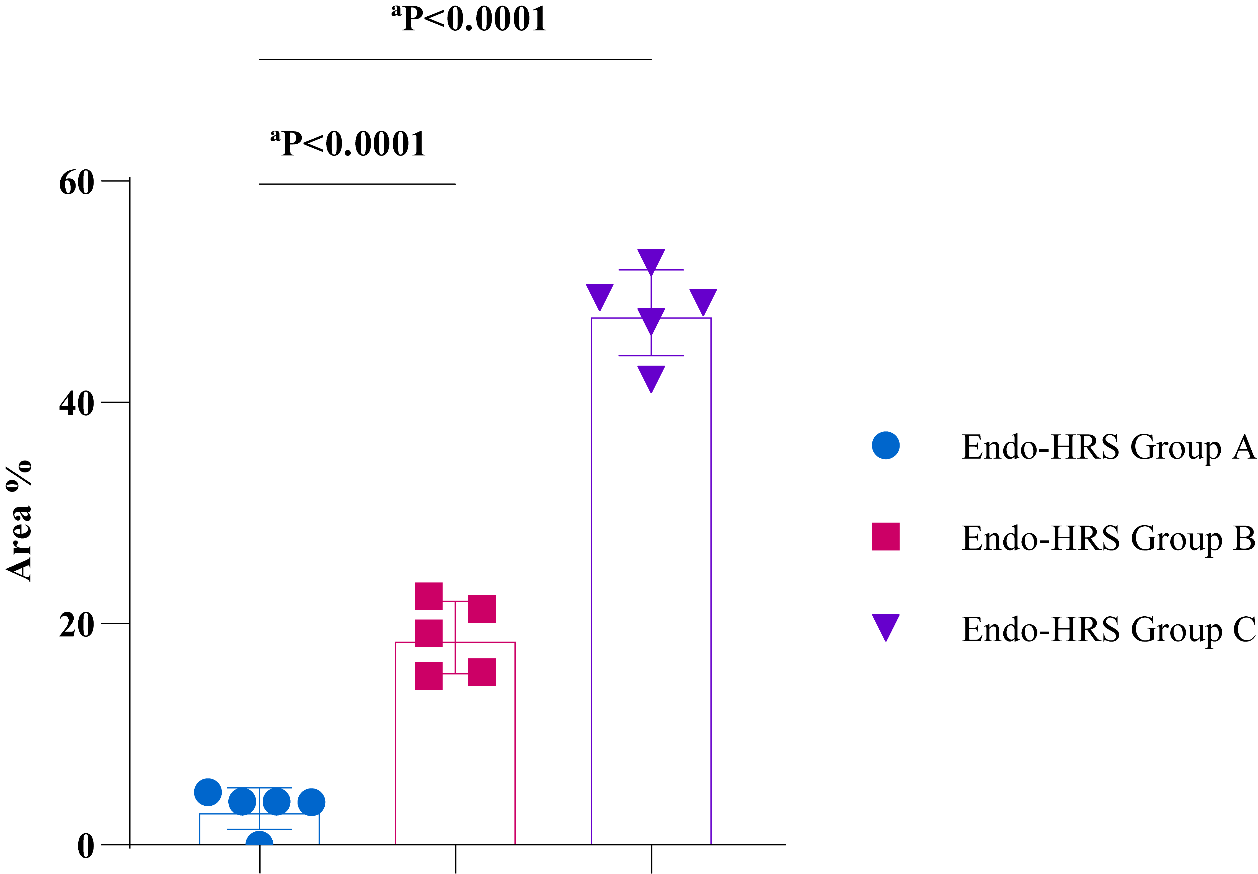
